# Variant location is a novel risk factor for individuals with arrhythmogenic cardiomyopathy due to a desmoplakin (*DSP*) truncating variant

**DOI:** 10.1101/2021.10.16.21264154

**Authors:** Edgar T. Hoorntje, Charlotte Burns, Luisa Marsili, Ben Corden, Victoria N. Parikh, Gerard J. te Meerman, Belinda Gray, Ahmet Adiyaman, Richard D. Bagnall, Daniela Q.C.M. Barge-Schaapveld, Maarten P. van den Berg, Marianne Bootsma, Laurens P. Bosman, Gemma Correnti, Johan Duflou, Ruben N. Eppinga, Diane Fatkin, Michael Fietz, Eric Haan, Jan D.H. Jongbloed, Arnaud D. Hauer, Lien Lam, Freyja H.M. van Lint, Amrit Lota, Carlo Marcelis, Hugh J. McCarthy, Anneke M. van Mil, Rogier A. Oldenburg, Nicholas Pachter, R. Nils Planken, Chloe Reuter, Christopher Semsarian, Jasper J. van der Smagt, Tina Thompson, Jitendra Vohra, Paul G.A. Volders, Jaap I. van Waning, Nicola Whiffin, Arthur van den Wijngaard, Ahmad S. Amin, Arthur A.M. Wilde, Gijs van Woerden, Laura Yeates, Dominica Zentner, Euan A. Ashley, Matthew T. Wheeler, James S. Ware, J. Peter van Tintelen, Jodie Ingles

**Author notes:** **ADDRESS FOR CORRESPONDENCE** Associate Professor Jodie Ingles, Centre for Population Genomics, Garvan Institute of Medical Research and Murdoch Children’s Research Institute, 384 Victoria Street, Darlinghurst NSW 2010 Australia, Twitter: @jodieingles27. These authors contributed equally (Edgar T. Hoorntje, Charlotte Burns, James S. Ware, J. Peter van Tintelen, Jodie Ingles). **ETHICS DECLARATION** Institutional ethics approval was granted by individual sites; including Sydney Local Health District, Royal Prince Alfred Hospital, Australia; Royal Brompton & Harefield Hospitals Cardiovascular Biobank (National Research Ethics Service), UK. Waiver of consent was granted at Stanford School of Medicine Internal Review Board, USA. All individual-level data were de-identified. **AUTHOR INFORMATION** 1. Conceptualization: CB, EJH, JW, JPvT, JI; 2. Data curation: ALL; 3. Formal Analysis: EJH, CB, JI; 4. Funding acquisition: JPvT, JI; 5. Investigation: EJH, CB, BC, VP, JW, PvT, JI; 6. Methodology: ALL; 6. Project administration: JI; 7. Resources: JW, PT, JI; 8. Software: Not Applicable; 9: Supervision: JW, PvT, JI; 10. Validation: Not Applicable; 11. Visualization: JI; 12. Writing – original draft: EJH, CB, JI; 13. Writing – review & editing: ALL.

## Abstract

**Background:** Truncating variants in desmoplakin (*DSP*tv) are an important cause of arrhythmogenic cardiomyopathy (ACM), however the genetic architecture and genotype-specific risk factors are incompletely understood. We evaluated phenotype, risk factors for ventricular arrhythmias, and underlying genetics of *DSP*tv cardiomyopathy.

**Methods:** Individuals with *DSP*tv and any cardiac phenotype, and their gene-positive family members were included from multiple international centers. Clinical data and family history information were collected. Event-free survival from ventricular arrhythmia was assessed. Variant location was compared between cases and controls, and literature review of reported *DSP*tv performed.

**Results:** There were 98 probands and 72 family members (mean age at diagnosis 43 ± 18 years, 59% female) with a *DSP*tv, of which 146 were considered clinically affected. Ventricular arrhythmia (sudden cardiac arrest, sustained ventricular tachycardia, appropriate implantable cardioverter defibrillator therapy) occurred in 56 (33%) individuals. *DSP*tv location and proband status were independent risk factors for ventricular arrhythmia, while prior risk factors showed no association. Further, gene region was important with variants in cases (cohort n=98, Clinvar n=168) more likely to occur in the regions resulting in nonsense mediated decay of both major *DSP* isoforms, compared to n=124 gnomAD control variants (148 [83.6%] versus 29 [16.4%], p<0.0001).

**Conclusions:** In the largest series of individuals with *DSP*tv, we demonstrate variant location is a novel risk factor for ventricular arrhythmia, can inform variant interpretation, and provide critical insights to allow precision-based clinical management.

## INTRODUCTION

Desmoplakin is a plakin family protein that anchors the desmosome to intermediate filaments and is abundant in tissues with greater mechanical stress such as the epidermis and myocardium (1,2). Genetic variants in the gene encoding desmoplakin (*DSP*) cause a range of cardio-cutaneous phenotypes including arrhythmogenic cardiomyopathy (ACM), striate palmoplantar keratoderma and lethal acantholytic epidermolysis bullosa in more severe cases (3). Truncating variants (*DSP*tv) that lead to putative loss of function (LOF) via haploinsufficiency of the protein have been previously reported as causative of disease (2). *DSP*-null mice show extensive disruption of the cytoarchitecture and cell resilience in skin and heart tissue, with death in early development (4).

Arrhythmogenic right ventricular cardiomyopathy (ARVC), the right dominant sub-form of ACM (2,5), is characterised by progressive loss and fibrofatty replacement of the ventricular myocardium (6). Diagnosis of ARVC can be challenging and 2010 Task Force Criteria consider electrical, structural (imaging and histological) and genetic characteristics (7). Historically, clinical descriptions of *DSP*tv were often based on ARVC cohorts, though growing recognition of left ventricular (LV) involvement has necessitated a shift to a broader phenotype description, ACM (8), encompassing left dominant arrhythmogenic cardiomyopathy (LDAC) and biventricular disease, with new Padua criteria proposed (9). Dilated cardiomyopathy (DCM) and LDAC lie on a spectrum, with overlap in molecular causes. The phenotypic label reflects whether rhythm disturbance, cardiac dilation, or contractile imparement predominates (10). In one of the largest studies to date, clinical characteristics of *DSP* variants in a population of 44 probands and 63 family members were reported as a distinct ACM characterised by LV fibrosis, myocardial inflammation and high incidence of ventricular arrhythmias (11). Biallelic *DSP* variants can give rise to Carvajal syndrome, characterised by woolly hair, palmoplantar keratoderma and development of ACM in childhood, and often due to homozygous or compound heterozygous *DSP*tv affecting the C-terminus (12).

The N-terminal globular head of *DSP* is important in desmosome organisation by binding plaque proteins such as plakophilin and plakoglobin, while the central rod domain contains a coiled-coil region (13). The C-terminal contains three plakin repeat domains, required for alignment and binding of intermediate filaments (14). Two predominant isoforms exist due to alternate splicing, *DSPI* which is the longest isoform and *DSPII* which has a shortened central rod domain. DSPI and DSPII are expressed in equivalent levels in epidermis, however DSPI is more prevalent in myocardium (15). Differences between the two isoforms relate to the rod domain size, considered important for self-association and formation of homo-dimers (16).

Here we report an international cohort of individuals with a *DSP*tv. We describe the phenotype spectrum of *DSP*tv cardiomyopathy, family history characteristics, and provide insights into the genetic architecture of *DSP*tv cardiomyopathy and its relation to clinical phenotype.

## METHODS

More detailed methods are available in the supplement.

### International cohort

An international (Australia, United Kingdom, Netherlands and United States of America) retrospective cohort of unrelated patients and family members with a *DSP*tv was assembled, comprising patients or their relatives seen in specialised cardiac genetic clinics, outpatient cardiology clinics or clinical genetics services. Cases were submitted between July 2016 and August 2018. All aspects of the study were performed according to institutional human research ethics committee approval according to the local sites.

### Eligibility criteria

*DSP*tv were those affecting a canonical splice-site, nonsense variants, or insertion or deletion variants leading to a frameshift. All variant nomenclature adhered to the Human Genome Variation Society sequence variant nomenclature recommendations, using reference transcript NM_004415.3. Probands and family members were included irrespective of cardiac phenotype, if they carried one of the variants described above. Both deceased and living patients were included, regardless of their age. Diagnosis was recorded by the referring institution and participants were classified as probands or family members, or as clinically affected (including any cardiac phenotype such as cardiomyopathy or ventricular arrhythmias, or cutaneous phenotype) and unaffected. Genetic testing was performed by the referring institution. More detailed information is available in the supplement.

### Variant location analyses

Genetic variant data were sought from three sources; (1) Variants identified in patients included in this study; (2) Variants submitted to ClinVar with a review status of one-star and above, that were classified as pathogenic or likely pathogenic (17) (Data downloaded on 5th October 2018); and (3) *DSP*tv listed in gnomAD v2.1 (18).

Variants were grouped in to different gene regions based on (i) whether the region is included in one or both of the 2 major isoforms *DSPI* and *DSPII* (**Figure 1**), and (ii) whether a variant in that location would be expected to trigger nonsense mediated decay (NMD). This results in three regions (**Figure 2**), a constitutive (incorporated in to both major isoforms) and NMD-competent region at the N-terminal (exons 1-22, part of exon 23, c.1-c.3582), a region that is non-constitutive (incorporated in to *DSPI* only) and NMD-competent located in the central rod domain (*DSPI/DSPIa* c.3583-c.4050 and *DSPI* c.4051-c.5379), and a region that is constitutive but NMD-incompetent at the C-terminus (exon 24; c.5324-8616).

**FIGURE 1:**
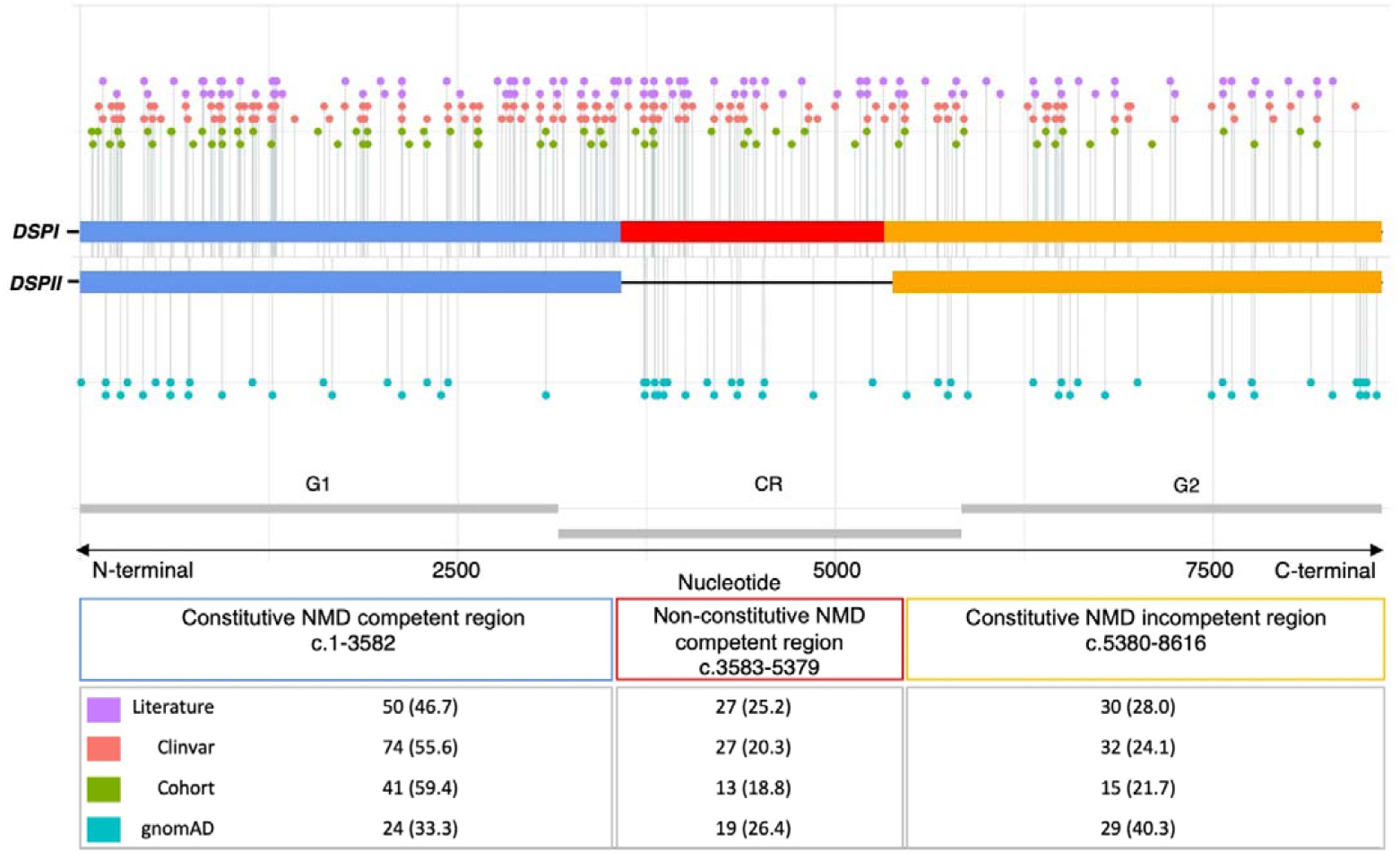
Linear topology schematic showing distribution of *DSP*tv across the key gene regions of *DSPI* and *DSPII*. Case variants (Cohort, Clinvar and Literature) are shown above and control variants below the line. Number of unique variants from each source are shown in the table. Abbreviation: NMD, nonsense mediated decay.

**FIGURE 2:**
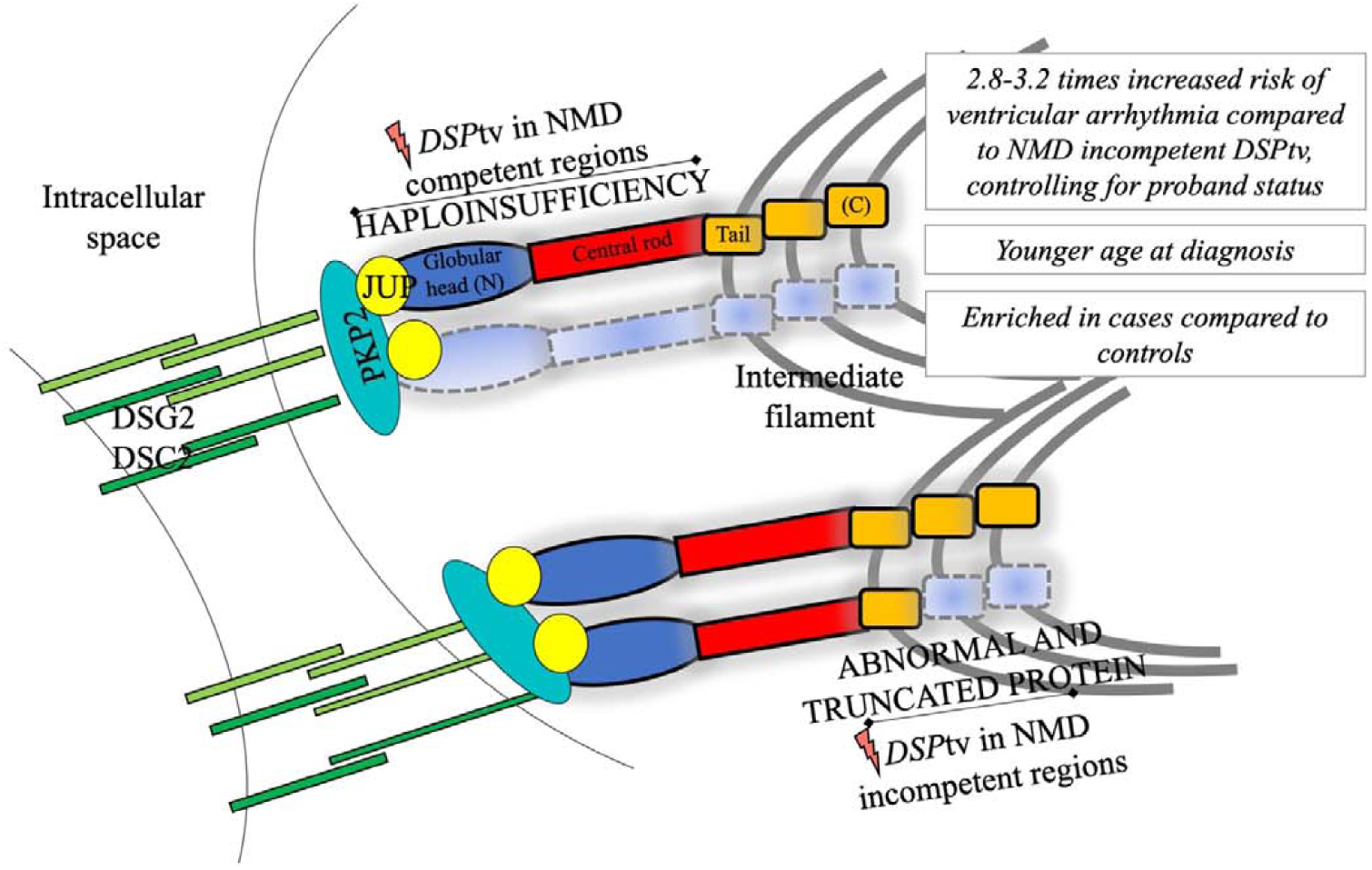
Summary of the key findings and illustration of the impact of *DSP*tv location on protein expression (for DSPI)

### Genetic variant classification

Disease variants were classified using the American College of Medical Genetics and Genomics and Association for Molecular Pathology (ACMG/AMP) standards for variant classification (19). In brief, the criterion pathogenic moderate (PM2) were allocated to variants with a frequency in the Genome Aggregation Database (gnomAD) of <0.04% (20). Pathogenic very strong (PVS1) was used for variants in the NMD-competent regions, while PVS1_strong for *DSP*tv in the constitutive NMD-incompetent region, with rationale for this provided in the supplement.

### Literature review

We conducted a literature review of all *DSP*tv reported in PubMed (accessed on June 3, 2019; Supplementary material).

### Statistical analysis

Data were analysed using RStudio (version 1.2). One-way analysis of variance or *t*-test was used for comparing continuous variables and chi-square or Fisher exact test for categorical variables. Multivariate Cox proportional hazards models were used to assess freedom from ventricular arrhythmia, using time since birth as the time variable. For patients with ventricular arrhythmia, time to first event was used. Where there was no ventricular arrhythmia, the date of the last known cardiac evaluation was used for censoring. Hazard ratios and 95% confidence interval (CI) were calculated. To account for family-clustering in the data, analyses were also repeated in a proband only dataset. Variables reaching p<0.05 in univariate Cox regression were included in the multivariate Cox regression model, using a backward step-wise approach.

## RESULTS

### Study population

Overall there were 98 probands (mean age at diagnosis 42 ± 18 years, 59% female) and 72 family members identified (mean age at diagnosis 45 ± 19 years, 61% female; **Table 1**). There were 95 probands with a cardiomyopathy and 3 with a primary cutaneous phenotype. Among family members, 48/72 were deemed affected, including 5 with a predominantly cutaneous phenotype. In total, 146 individuals were considered affected including cardiomyopathy, ventricular arrhythmia and cutaneous phenotypes.

**TABLE 1:** Clinical characteristics by proband status and sex.

|  |  | **Proband status** |  | **Sex (Affected only)** |  |  |
| --- | --- | --- | --- | --- | --- | --- |
| **Diagnosis** | **Total**<br>**n=178** | **Probands**<br>**n=98** | **Family members**<br>**n=72** | **Female**<br>**n=86** | **Male**<br>**n=60** | **p-value** |
| Affected | 146 | 98 (100) | 48 (67) | 86 (100) | 60 (100) | NA |
| Mean age at diagnosis ± SD, years | 43 ± 18 | 42 ± 18 | 45 ± 19 | 40 ± 17 | 46 ± 19 | 0.07 |
| Female sex | 101 (59) | 57 (58) | 44 (61) | NA | NA | NA |
| T wave inversion beyond V3 | 33 (24) | 25 (32) | 8 (13) | 22 (33) | 9 (18) | 0.07 |
| Low voltages | 47 (35) | 30 (39) | 17 (29) | 28 (42) | 13 (27) | 0.09 |
| Premature ventricular contractions | 35 (25) | 23 (29) | 12 (20) | 25 (35) | 10 (21) | 0.09 |
| Ventricular arrhythmia | 56 (33) | 46 (47) | 10 (14) | 37 (44) | 19 (32) | 0.17 |
| Sudden cardiac death | 13 (8) | 10 (10) | 3 (4) | 7 (8) | 6 (10) | 0.71 |
| Resuscitated cardiac arrest | 15 (9) | 12 (12) | 3 (20) | 8 (10) | 7 (12) | 0.65 |
| Sustained VT | 19 (11) | 15 (15) | 4 (6) | 14 (17) | 5 (8) | 0.24 |
| Appropriate ICD therapy | 16 (10) | 16 (16) | 0 (0) | 13 (15) | 3 (5) | 0.09 |
| End-stage heart failure | 10 (8) | 7 (8) | 3 (5) | 6 (8) | 4 (8) | 0.94 |
| Heart transplant / LVAD | 6 (6) | 6 (6) | 0 (0) | 4 (5) | 2 (3) | 0.64 |
| Combined endpoint | 64 (38) | 52 (53) | 12 (17) | 41 (48) | 23 (39) | 0.27 |
| Myocarditis | 8 (10) | 6 (13) | 2 (6) | 2 (5) | 6 (24) | 0.05 |
| **Transthoracic echocardiogram** |  |  |  |  |  |  |
| LV ejection fraction | 40 ± 16 | 35 ± 15 | 48 ± 13 | 35 ± 14 | 42 ± 15 | 0.03 |
| LV end diastolic diameter | 57 ± 9 | 59 ± 9 | 53 ± 8 | 58 ± 8 | 59 ± 10 | 0.56 |
| LV end systolic diameter | 44 ± 12 | 47 ± 12 | 37 ± 9 | 47 ± 12 | 44 ± 12 | 0.29 |
| **CMR imaging** |  |  |  |  |  |  |
| LV ejection fraction, % | 42 ± 12 | 38 ± 12 | 48 ± 12 | 39 ± 11 | 43 ± 13 | 0.14 |
| LV end diastolic volume, indexed | 111 ± 33 | 117 ± 35 | 100 ± 23 | 119 ± 34 | 105 ± 29 | 0.09 |
| LV end systolic volume, indexed | 67 ± 32 | 73 ± 35 | 54 ± 20 | 74 ± 34 | 60 ± 27 | 0.07 |
| RV ejection fraction, % | 47 ± 12 | 44 ± 12 | 53 ± 8 | 47 ± 11 | 45 ± 12 | 0.46 |
| RV end diastolic volume, indexed | 89 ± 24 | 93 ± 25 | 83 ± 21 | 84 ± 20 | 100 ± 27 | 0.01 |
| RV end systolic volume, indexed | 48 ± 19 | 52 ± 19 | 41 ± 16 | 46 ± 17 | 56 ± 20 | 0.05 |
| Regional wall motion abnormalities | 44 (45) | 35 (56) | 9 (26) | 25 (46) | 19 (51) | 0.64 |
| Late gadolinium enhancement | 59 (61) | 40 (63) | 19 (58) | 31 (57) | 28 (76) | 0.07 |
| LV noncompaction | 22 (22) | 17 (27) | 5 (13) | 14 (29) | 8 (21) | 0.42 |

Data shown are n (%) or mean ± standard deviation. Abbreviations: ICD, implantable cardioverter defibrillator; LVAD, left ventricular assist device; VT, ventricular tachycardia; LV, left ventricular; CMR, cardiac magnetic resonance; RV, right ventricle.

### Sex differences

Females were over-represented compared to males among affected individuals (86 [59%] versus 60 [41%]; **Table 1**). There was no difference in mean age at diagnosis between females and males (40 ± 17 years versus 46 ± 19 years, p=0.07). Myocarditis was reported more frequently in males (2/43 [5%] versus 6/25 [24%], p<0.05) but this was not always reliably reported. Women had reduced LV ejection fraction (LVEF) on transthoracic echocardiography, but not cardiac magnetic resonance imaging (CMR) derived LVEF. Men had greater indexed RV end diastolic volume (84 ± 20 versus 100 ± 27, p=0.01). No difference in clinical outcomes were reported between sexes. Women had comparable distribution of variants by gene region compared to men.

### Electrophysiological characteristics

There was a high rate of ventricular arrhythmia occurring in 56 (33%) individuals, including 46 (47%) probands and 10 (14%) family members. Ventricular arrhythmia included sudden cardiac death (SCD) in 13 (8%; 10 probands), resuscitated cardiac arrest in 15 (9%; 12 probands), appropriate implantable cardioverter defibrillator (ICD) therapy in 16 (10%; 16 probands) and sustained ventricular tachycardia in 19 patients (11%; 15 probands); including 10 (14%) family members, and with some experiencing multiple events. Six probands experienced two ventricular arrhythmia episodes, initially having sustained ventricular tachycardia (n=4) or resuscitated cardiac arrest (n=2), followed by appropriate ICD therapy. SCD or resuscitated cardiac arrest was the presenting symptom in 24 (14%; 20 probands) patients. T wave inversion beyond V3 occurred in 33 (24%), low voltages in 47 (35%) and premature ventricular contractions in 56 (33%).

### Imaging characteristics

Echocardiographic and CMR characteristics are shown in **Table 1**. Signs of LV noncompaction (LVNC) were reported (n=22), with 6 having a ratio of noncompacted to compacted layer >2.3 on CMR. Four probands were reported to have hypertrophic cardiomyopathy (HCM) with ages at diagnosis ranging from 58-83 years, and LV hypertrophy measuring 26mm, 16mm, and an apical pattern in two. *DSP*tv are not established as associated with HCM, and we consider it unlikely that these variants are causal for HCM for these 4 cases, but are reported as they met the pre-specified eligibility criteria. Late gadolinium enhancement (LGE) was reported in 59 (61%), and end-stage heart failure was reported in 10 (8%) patients. Two females developed disease while pregnant, one showed impaired LV function (LVEF <45%) at 32 weeks of gestation, while the other developed narrow complex tachycardia at 38 weeks of gestation with subsequent echocardiogram showing a dilated and impaired LV. A further two women developed disease during the postpartum period. Finally, another patient who died suddenly during pregnancy was identified to be positive for Parvovirus B19 on postmortem Parvo-polymerase chain reaction in myocardial tissue. Myocarditis was reported in 7 individuals on CMR (and another on postmortem investigation).

### Genetic analysis

A total of 69 distinct *DSP*tv were identified in the 98 probands (**Supplementary Table 1**). Among the 69 *DSP*tv, there were 31 small insertions or deletions leading to a frameshift and downstream premature termination codon, 25 nonsense variants, 12 canonical splice-site altering variants and a large deletion of exons 5-24. Eleven (16%) variants were classified as pathogenic, 57 (83%) were classified as likely pathogenic and 1 (1%) was classified as a variant of uncertain significance. Two probands had a diagnosis of cardiomyopathy, with woolly hair and keratoderma (OMIM 605676) and were compound heterozygous, each carrying a *DSP*tv (p.Arg2229Serfs*32 or p.Tyr28Alafs*66) and a *DSP* splice site variant (the same c.273+5G>A in both). This splice site variant has an allele count of 79 in gnomAD, with an allele frequency of 0.028% and considered a variant of uncertain significance under a recessive inheritance model.

### *DSP*tv location

We investigated whether case and control variants localised to the specified gene regions, constitutive NMD-competent, non-constitutive NMD-competent and constitutive NMD-incompetent (**Figure 1**). Pathogenic and likely pathogenic *DSP*tv submitted to ClinVar, as well as variants described above in the international cohort were included, giving a total of 266 cases. This included 69 unique *DSP*tv identified in 98 individuals in the international cohort and 135 unique *DSP*tv from 168 cases reported in ClinVar (**Supplementary Table 2**). One variant reported in ClinVar was excluded from analysis given it resided in the small overlap region of exon 23 which is both non-constitutive and predicted NMD-incompetent due to being <55 bp upstream of the last exon junction (*DSP*: c.5327_5330del, p.Glu1776Glyfs). Literature cases are shown in **Figure 1** but not included in the analysis due to unquantified sample overlap. Variants observed in cases were compared to 72 unique *DSP*tv observed as 124 alleles in gnomAD controls (**Supplementary Table 3**). Case variants were more frequently seen in the constitutive NMD-competent region compared to controls. Across the 3 gene regions (constitutive NMD-competent, non-constitutive NMD-competent and constitutive NMD-incompetent, respectively), *DSP*tv were seen in 148 (56%), 59 (22%) and 59 (22%) cases compared to *DSP*tv observed in controls 29 (23%), 28 (23%) and 67 (54%), overall p<0.001.

Clinical characteristics of patients with *DSP*tv in the three gene regions are shown in **Table 2**. Overall there were few significant differences between the patient groups based on gene region. Age at diagnosis was significantly younger in those with *DSP*tv in both NMD-competent regions (constitutive and non-constitutive). Further, there was a greater risk of ventricular arrhythmia and risk of the combined endpoint in those with *DSP*tv in the constitutive and non-constitutive NMD-competent regions.

**TABLE 2:** Cardiac investigation of affected individuals with *DSP*tv by gene region.

|  | N | Constitutive<br>NMD-competent | Non-constitutive<br>NMD-competent | Constitutive<br>NMD-incompetent | p-value |
| --- | --- | --- | --- | --- | --- |
| n | 146 | 83 (57) | 36 (25) | 27 (18) | - |
| Age at diagnosis, years | 143 | 40 ± 18 | 37 ± 16 | 52 ± 16 | 0.002 |
| Female sex | 146 | 49 (59) | 17 (63) | 20 (56) | 0.84 |
| Family history of disease | 107 | 36 (57) | 15 (68) | 9 (43) | 0.25 |
| Ventricular arrhythmia | 144 | 34 (41) | 14 (54) | 8 (22) | 0.03 |
| SCD | 143 | 7 (9) | 4 (15) | 2 (6) | 0.40 |
| Resuscitated cardiac arrest | 143 | 7 (9) | 5 (23) | 2 (6) | 0.06 |
| Sustained VT | 143 | 14 (17) | 1 (4) | 4 (11) | 0.19 |
| Appropriate ICD therapy | 143 | 13 (16) | 3 (12) | 0 (0) | 0.04 |
| Combined endpoint | 144 | 39 (48) | 15 (58) | 10 (28) | 0.04 |
| Myocarditis | 68 | 6 (18) | 1 (5) | 1 (7) | 0.32 |
| LV noncompaction | 87 | 14 (26) | 6 (40) | 2 (11) | 0.16 |
| Premature ventricular contractions | 119 | 21 (33) | 4 (18) | 10 (30) | 0.43 |
| **Transthoracic echocardiogram** |  |  |  |  |  |
| LV ejection fraction | 100 | 41 ± 16 | 28 ± 11 | 38 ± 13 | 0.008 |
| LV end diastolic diameter | 98 | 59 ± 9 | 57 ± 10 | 57 ± 7 | 0.71 |
| LV end systolic diameter | 77 | 45 ± 12 | 47 ± 14 | 46 ± 9 | 0.95 |
| **CMR imaging** |  |  |  |  |  |
| LV ejection fraction | 88 | 41 ± 12 | 38 ± 13 | 41 ± 11 | 0.55 |
| LV end diastolic volume, indexed | 75 | 121 ± 39 | 114 ± 28 | 104 ± 22 | 0.13 |
| LV end systolic volume, indexed | 76 | 75 ± 37 | 69 ± 30 | 61 ± 24 | 0.27 |
| RV ejection fraction | 62 | 49 ± 10 | 45 ± 11 | 44 ± 14 | 0.35 |
| RV end diastolic volume, indexed | 56 | 92 ± 26 | 81 ± 14 | 93 ± 25 | 0.35 |
| RV end systolic volume, indexed | 55 | 51 ± 19 | 48 ± 13 | 51 ± 21 | 0.87 |
| Late gadolinium enhancement | 91 | 29 (64) | 12 (63) | 18 (67) | 0.97 |
| Regional wall motion abnormalities | 91 | 22 (47) | 6 (35) | 16 (59) | 0.29 |
| **Electrocardiogram** |  |  |  |  |  |
| Sinus rhythm | 81 | 43 (98) | 16 (100) | 21 (100) | 0.65 |
| PR interval | 110 | 167 ± 43 | 155 ± 27 | 172 ± 50 | 0.35 |
| QRS | 108 | 102 ± 17 | 95 ± 12 | 97 ± 16 | 0.21 |
| QTc | 105 | 424 ± 40 | 417 ± 32 | 421 ± 39 | 0.76 |
| T wave inversion beyond V3 | 117 | 21 (33) | 3 (13) | 7 (23) | 0.14 |
| Low voltages | 114 | 23 (37) | 8 (38) | 10 (33) | 0.93 |

Data shown are mean ± standard deviation (n = number of persons with available data) or n (%). Abbreviations: LV, left ventricle; RV, right ventricle; NMD, nonsense mediated decay; SCD, sudden cardiac death; ICD, implantable cardioverter defibrillator.
Data shown are mean ± standard deviation or n (%)

### Event-free survival from ventricular arrhythmia based on gene region

Information with regard to occurrence of ventricular arrhythmia or censoring was available for 167 individuals. There were 56 probands and family members who experienced a ventricular arrhythmia during their lifetime. Univariate Cox proportional hazards models showed gene region and proband status as significantly associated with worse survival from ventricular arrhythmias (**Table 3; Figure 3**). Adjusting for other variables, variants in the constitutive NMD competent region (HR 2.8, 95% CI 1.3-6.0, p=0.01), non-constitutive NMD-competent region (HR 3.2, 95%CI 1.3-7.9, p=0.0009) and proband status (HR 3.3, 95%CI 1.7-6.6, p=0.0006) remained significant independent life-time risk factors for ventricular arrhythmia (**Table 3**).

**FIGURE 3:**
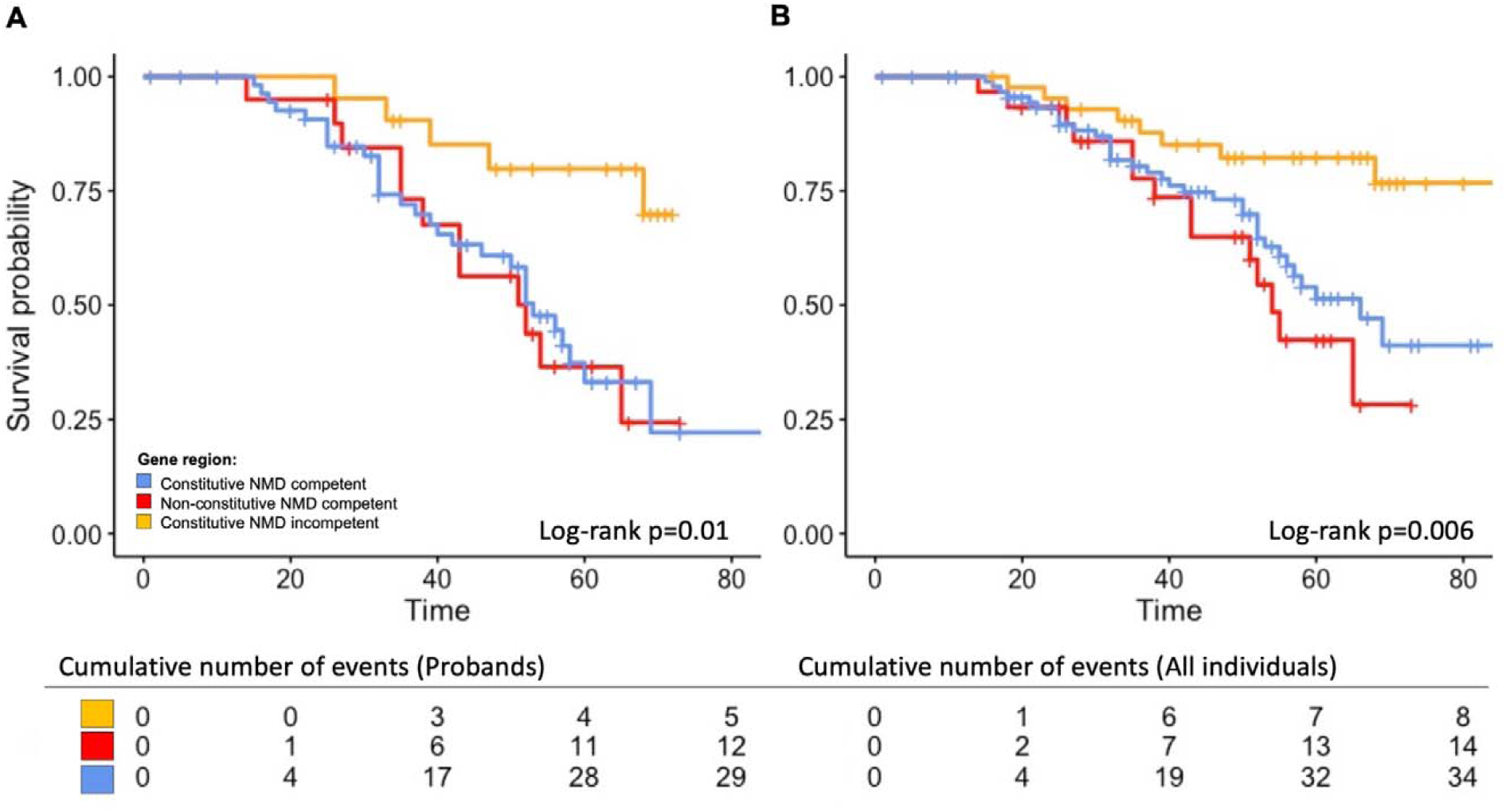
Independent life-time risk factor for ventricular arrhythmia. (A) Gene region including probands only, (B) Gene region including probands and affected family members. Time, age in years. Abbreviation: NMD, nonsense mediated decay.

**TABLE 3:**
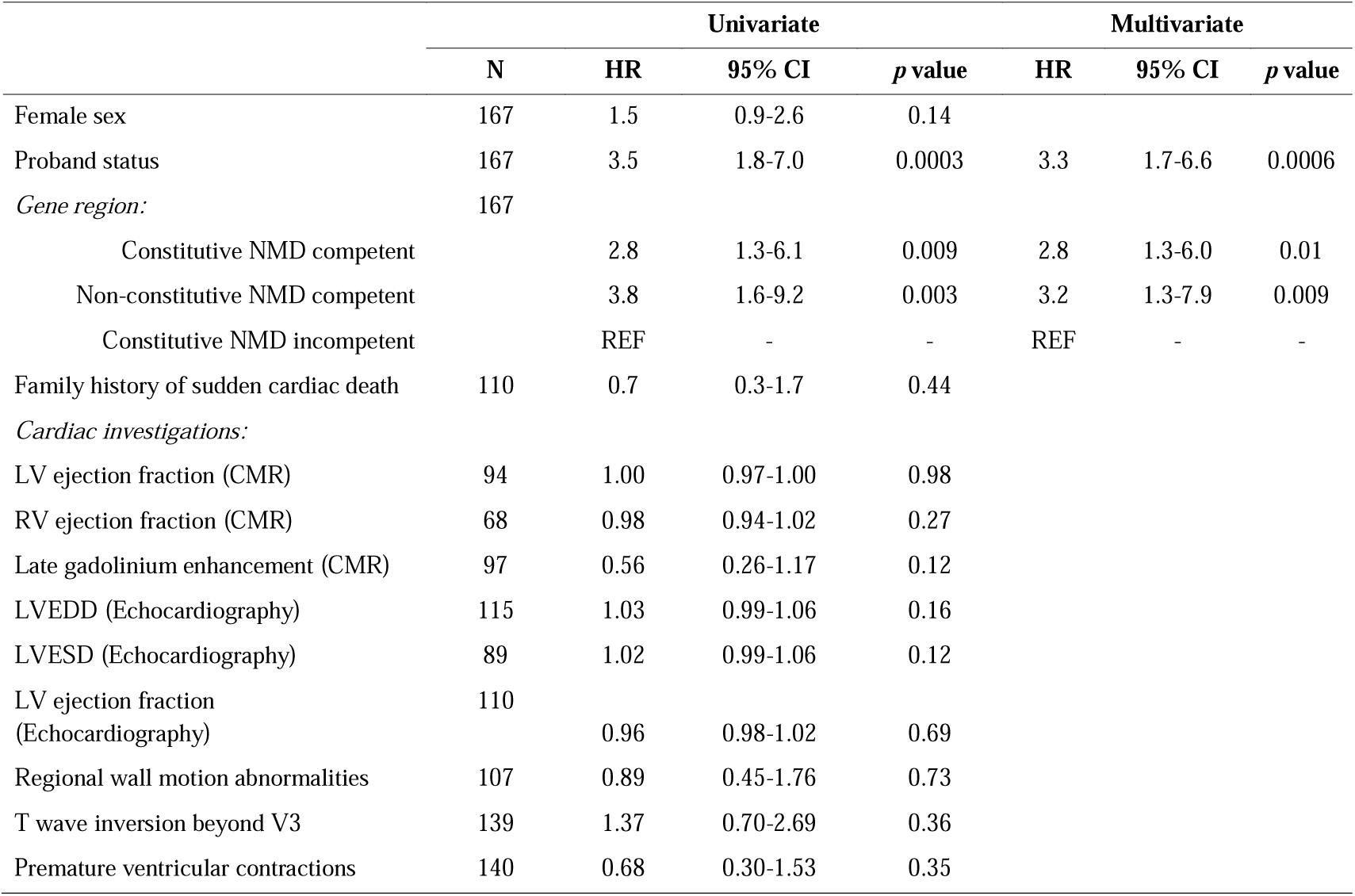
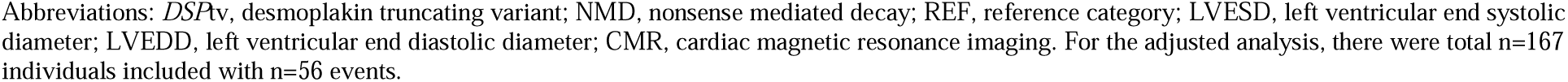
Life time risk factors for ventricular arrhythmia for individuals with a *DSP*tv.

|  | Univariate |  |  |  | Multivariate |  |  |
| --- | --- | --- | --- | --- | --- | --- | --- |
|  | N | HR | 95% CI | *p* value | HR | 95% CI | *p* value |
| Female sex | 167 | 1.5 | 0.9-2.6 | 0.14 |  |  |  |
| Proband status | 167 | 3.5 | 1.8-7.0 | 0.0003 | 3.3 | 1.7-6.6 | 0.0006 |
| *Gene region:* | 167 |  |  |  |  |  |  |
| Constitutive NMD competent |  | 2.8 | 1.3-6.1 | 0.009 | 2.8 | 1.3-6.0 | 0.01 |
| Non-constitutive NMD competent |  | 3.8 | 1.6-9.2 | 0.003 | 3.2 | 1.3-7.9 | 0.009 |
| Constitutive NMD incompetent |  | REF | - | - | REF | - | - |
| Family history of sudden cardiac death | 110 | 0.7 | 0.3-1.7 | 0.44 |  |  |  |
| *Cardiac investigations:* |  |  |  |  |  |  |  |
| LV ejection fraction (CMR) | 94 | 1.00 | 0.97-1.00 | 0.98 |  |  |  |
| RV ejection fraction (CMR) | 68 | 0.98 | 0.94-1.02 | 0.27 |  |  |  |
| Late gadolinium enhancement (CMR) | 97 | 0.56 | 0.26-1.17 | 0.12 |  |  |  |
| LVEDD (Echocardiography) | 115 | 1.03 | 0.99-1.06 | 0.16 |  |  |  |
| LVESD (Echocardiography) | 89 | 1.02 | 0.99-1.06 | 0.12 |  |  |  |
| LV ejection fraction (Echocardiography) | 110 | 0.96 | 0.98-1.02 | 0.69 |  |  |  |
| Regional wall motion abnormalities | 107 | 0.89 | 0.45-1.76 | 0.73 |  |  |  |
| T wave inversion beyond V3 | 139 | 1.37 | 0.70-2.69 | 0.36 |  |  |  |
| Premature ventricular contractions | 140 | 0.68 | 0.30-1.53 | 0.35 |  |  |  |

Abbreviations: *DSP*tv, desmoplakin truncating variant; NMD, nonsense mediated decay; REF, reference category; LVESD, left ventricular end systolic diameter; LVEDD, left ventricular end diastolic diameter; CMR, cardiac magnetic resonance imaging. For the adjusted analysis, there were total n=167 individuals included with n=56 events.

### Cutaneous phenotype

Cutaneous abnormalities were not systematically reported, however notably one family with a *DSP*tv in the non-constitutive NMD-competent region (cardiac isoform, *DSPI*) had an affected relative with hyperkeratosis and cardiomyopathy. An additional 13 individuals with *DSP*tv in the constitutive NMD-competent region and 5 in the constitutive NMD-incompetent region were reported with overt cardio-cutaneous features noted at clinical review. In eight patients (5%; 3 probands) only cutaneous abnormalities were reported, and were the sole finding in one family following an autosomal dominant inheritance pattern.

### Postmortem findings and cardiac transplant histology

Thirteen patients (8%; 10 probands) presented with SCD (**Supplementary Table 4**). In all 13, a postmortem investigation was performed. The mean age at death was 26 ± 11 years. Where recorded, the activity at time of death varied from exercise through to sleep. No decedent had a pre-morbid diagnosis of a cardiac condition. Nine decedents received a postmortem diagnosis of ARVC or probable ARVC. There was LV involvement in all cases and fibrosis and fatty infiltration commonly reported.

Two patients underwent a heart transplant due to end stage heart failure. Biventricular involvement was observed in both hearts, as were signs of LVNC. One heart showed ARVC with septal involvement and replacement fibrosis in both ventricles and septum. The other heart showed LVNC with notable RV involvement consisting of fatty changes and atrophy.

### Family history characteristics

Among the probands, 49 (51%) had a documented family history of cardiomyopathy, while 16 (17%) had a family history of a suspicious SCD under the age of 40 years. Of 72 family members with positive gene results included, 48 (67%) had overt disease, while 24 (33%) remained asymptomatic (mean age of 49 ± 22 years and 15 [63%] were female). There were 8 family members aged 60 years or older (60-86 years; 5 females) with no clinical evidence of disease, suggesting incomplete penetrance. By gene region, there was no statistical difference in the proportion of probands with a positive family history (constitutive NMD-competent 27 [49%], non-constitutive NMD-competent 13 [65%], constitutive NMD-incompetent 9 [43%], p=0.33).

### Literature review of reported *DSP*tv

Three hundred and fifteen studies were identified, 240 were screened and 185 full texts were assessed for eligibility (85 were excluded from the final qualitative synthesis, including 66 that did not report any *DSP* variant, 2 where phenotype was not provided, 2 with no full text article available, and 1 review; **Supplementary Figure 1**). Of 98 studies included in the final selection, a total of 105 *DSP*tv in 143 probands from apparently unrelated families were reported, including 57 nonsense, 42 frameshift, and 6 splice site variants (**Supplementary Tables 5-7**). All reported variants were absent or very rare (allele count ≤ 2) in gnomAD and were classified as pathogenic or likely pathogenic. One variant (p.Thr2104fs*12) was present 13 times in gnomAD however has strong evidence of pathogenicity and reported in a compound heterozygous state.

Both dominant and recessive patterns of inheritance of *DSP*tv were reported. Cascade genetic testing to confirm autosomal dominant inheritance was reported for only 19 *DSP*tv (dominant *DSP*tv) in 22 families. Families reported with autosomal dominant inheritance commonly demonstrated adult age of onset, incomplete penetrance and variable clinical expression. Of 105 reported *DSP*tv, 26 were only identified in affected individuals with homozygous or compound heterozygous inheritance. Four *DSP*tv co-occurred in trans with one of three missense *DSP* variants (p.Ala2655Asp, p.Arg2366Cys and p.Asn287Lys), each of which involved highly conserved residues within globular heads, are absent in gnomAD, and classified as likely pathogenic. There were 16 individuals with 23 *DSP*tv identified to have autosomal recessive disease, either homozygous (n=9) or compound heterozygous (n=7). In just those variants identified in a homozygous state there was only 1 (11%) in the constitutive NMD-competent region, 5 (56%) in the non-constitutive NMD-competent region and 3 (33%) in the constitutive NMD-incompetent region.

## DISCUSSION

*DSP*tv lead to a distinct cardiomyopathy characterised by LV involvement and a high-risk of ventricular arrhythmia and SCD. We present a large international series of cases with *DSP*tv and demonstrate that the precise location of the *DSP*tv is a novel risk factor for ventricular arrhythmia (**Figure 3**). Truncating variants in the constitutive NMD-competent region were enriched in cases compared to controls, and predicted to result in NMD and haploinsufficiency of both DSPI and DSPII. Established risk factors were not associated with ventricular arrhythmias in our population, though note our study population comprised many historical cases and those presenting with sudden cardiac arrest where pre-event data were incomplete. Nevertheless, our findings highlight the importance of personalized medicine and the move towards gene-guided management of patients in the future.

### Arrhythmogenic cardiomyopathy phenotype

Ventricular arrhythmias occur frequently in patients with *DSP*tv cardiomyopathy, with one previous study reporting 23% presenting with SCD events (21). In our cohort, 47% of probands had ventricular arrhythmia either at presentation or during follow-up. In addition, 14% of relatives experienced ventricular arrhythmia, including 7% as their initial presenting symptom. This included two probands who presented with resuscitated cardiac arrest without any overt structural abnormalities of the heart, supporting the notion that life-threatening electrical phenotype can precede overt cardiac structural disease (22-24). Our findings do not align with previously described risk factors for ventricular arrhythmia in ARVC and ACM. A recent series of 107 patients with any *DSP* variants (n=30 events) showed ventricular arrhythmias were associated with reduced LVEF (11). This finding was not replicated in our cohort, indeed LVEF <35% was present in nearly half of those with/without ventricular arrhythmia. Missing data were evaluated to determine if this led to an ascertainment bias in our cohort, however LVEF was complete for 40/56 (71%) and 92/112 (82%) with/without ventricular arrhythmia respectively, suggesting this is unlikely. In this same study, premature ventricular contractions (>500 beats in 24 hours), LGE and RV dysfunction were likewise not shown to be associated with ventricular arrhythmia. A limitation of our work is the lack of systematic evaluation and quantification of LGE, which might have shown an association with outcomes. In ARVC more broadly, the recent ARVC risk calculator showed inverted T waves in the precordial and inferior leads was an independent risk factor for events (25). Finally, we show family history of SCD, which has not previously been evaluated in this group, is not associated with ventricular arrhythmia in our population.

Prior observation that *DSP*tv are predominantly associated with a left dominant form of ACM (2,5,26) is in line with our findings. Females were overrepresented in our population, but otherwise shared similar clinical characteristics compared to males, except reduced indexed RV end diastolic diameter on CMR. This finding is in contrast to other reported inherited cardiomyopathy patient cohorts, where a higher prevalence of males is often reported (27-29). On ECG, *DSP*tv cardiomyopathy patients frequently had low QRS voltage and negative T waves beyond V3. Low QRS voltage in limb leads have previously been shown to be associated with the presence and amount of LGE in a study of 79 patients with ARVC (43). Regional wall motion abnormalities on CMR and epicardial to mid wall LGE patterns in the LV were frequently seen in our cohort. Septal LGE frequently occurs in patients with LDAC (14), and recent work has shown patients with *DSP* and *FLNC* ACM are more likely to have LGE, often with a ring-like pattern, compared to other DCM genotypes (30). Four probands were reported to have HCM, however it should be noted that all 4 probands were male, presenting in older age and 3 had mild LV hypertrophy, all characteristics previously described in the non-familial sub-group of HCM (31). Previous assessment of the clinical validity of *DSP* variants causing HCM failed to identify sufficient evidence of gene-disease association (32). While our finding remains unclear, it seems reasonable to consider these clinical diagnoses as unrelated to the *DSP*tv.

### Cutaneous phenotype

A recent systematic evaluation of cutaneous abnormalities among *DSP*tv showed all patients expressed some degree of skin or hair abnormalities, except those with *DSP*tv in the non-constitutive NMD-competent region (cardiac isoform, DSPI) (33). Interestingly, we report one proband and their affected relative with palmoplantar keratoderma, with a *DSP*tv in the non-constitutive NMD-competent region. Another study reported 10% of *DSP*tv had cutaneous disease only, while 12% were reported to have LV dominant ACM and cutaneous disease (5). There is strong evidence of the need to systematically screen for subtle cutaneous abnormalities in all *DSP*tv families.

### Genotype

We show *DSP*tv localised to the constitutive NMD-competent region, corresponding to the N-terminal globular head, were enriched in patients compared to controls, and this finding was replicated in the variants identified through literature review. This region plays a critical role in organisation and assembly of the desmosomal complex by binding with plakophilin and plakoglobin. One previous report of *DSP* missense variants in patients with a clinical diagnosis of ARVC suggested a potential ‘hotspot’ N-terminal region, with 8/17 (47%) missense variants localised to the N-terminal compared to 1/28 (4%) of controls (p<0.0008) (34). Further, they concluded *DSP*tv were significantly more prevalent in ARVC cases than controls. Indeed, a recent study also showed clustering of missense variants in the N-terminal, but reported *DSP*tv to be more evenly distributed across the gene (11), potentially limited by sample size. While it seems likely that truncating variants in the NMD-incompetent region escape NMD and have a less deleterious impact, functional work to date has shown highly variable pattern of protein expression representing both haploinsufficiency and dominant negative effects (35). Our literature review identified biallelic *DSP*tv localized more often to the constitutive NMD-incompetent region compared to dominant *DSP*tv, suggesting that single heterozygous *DSP*tv are more likely to cause disease when occurring in the NMD-competent regions. Further, very few cases with homozygous variants in the constitutive NMD-competent region have been reported, with one example of a sib pair with severe lethal acantholytic epidermolysis bullosa, who died at 1 and 3 days respectively (36). It seems unlikely these infants were DSP null, given *DSP* knockout mice show embryonic lethality (4), suggesting expression of low-level truncated protein may be able to rescue the phenotype to some degree. Taken together, identification of a *DSP*tv in the NMD-competent regions should be considered important and may prompt gene and disease-specific adaptation and use of the ACMG/AMP criteria (19). We suggest *DSP*tv in this region be allocated very strong level of evidence, PVS1, when considering pathogenicity, when seen in an individual with a well characterised and concordant phenotype.

### Study Limitations

This was a large retrospective cohort study, and while it was an international effort, differences in practices and data collection by site meant some variables were incomplete. Furthermore, the event rate and data missingness precluded more detailed risk factor analyses. Diagnosis was made by the referring clinician and most recruitment was from specialised tertiary referral centres and therefore likely represents more severe phenotypes. The literature review was limited by publication bias, and inconsistent reporting of clinical, family and genetic information.

## Conclusion

We present a large international series of individuals with *DSP*tv and show gene region is a novel risk factor, specifically *DSP*tv leading to predicted NMD of truncated protein and haploinsufficiency of DSPI and/or DSPII is an independent risk factor for ventricular arrhythmias. By sub-typing disease by genotype there is increasing ability to offer precision medicine-based advice and therapies, and thereby improved outcomes for patients and their families.

## Data Availability

Data available pending request to the corresponding author and in accordance with site data sharing permissions.

## REFERENCES

1. Corrado D, Basso C, Pilichou K, Thiene G. Molecular biology and clinical management of arrhythmogenic right ventricular cardiomyopathy/dysplasia. Heart 2011;97:530–9.

2. Castelletti S, Vischer AS, Syrris P et al. Desmoplakin missense and non-missense mutations in arrhythmogenic right ventricular cardiomyopathy: Genotype-phenotype correlation. Int J Cardiol 2017;249:268–273.

3. Bauce B, Basso C, Rampazzo A et al. Clinical profile of four families with arrhythmogenic right ventricular cardiomyopathy caused by dominant desmoplakin mutations. Eur Heart J 2005;26:1666–75.

4. Gallicano GI, Kouklis P, Bauer C et al. Desmoplakin is required early in development for assembly of desmosomes and cytoskeletal linkage. J Cell Biol 1998;143:2009–22.

5. Lopez-Ayala JM, Gomez-Milanes I, Sanchez Munoz JJ et al. Desmoplakin truncations and arrhythmogenic left ventricular cardiomyopathy: characterizing a phenotype. Europace. 2014;16:1838–46.

6. Hoorntje ET, Te Rijdt WP, James CA et al. Arrhythmogenic cardiomyopathy: pathology, genetics, and concepts in pathogenesis. Cardiovasc Res 2017;113:1521–1531.

7. Marcus FI, McKenna WJ, Sherrill D et al. Diagnosis of arrhythmogenic right ventricular cardiomyopathy/dysplasia: proposed modification of the task force criteria. Circulation 2010;121:1533–41.

8. Corrado D, van Tintelen PJ, McKenna WJ et al. Arrhythmogenic right ventricular cardiomyopathy: evaluation of the current diagnostic criteria and differential diagnosis. Eur Heart J 2020;41:1414–1429.

9. Corrado D, Perazzolo Marra M, Zorzi A et al. Diagnosis of arrhythmogenic cardiomyopathy: The Padua criteria. Int J Cardiol 2020;319:106–114.

10. Sen-Chowdhry S, Syrris P, Prasad SK et al. Left-dominant arrhythmogenic cardiomyopathy: an under-recognized clinical entity. J Am Coll Cardiol 2008;52:2175–87.

11. Smith ED, Lakdawala NK, Papoutsidakis N et al. Desmoplakin Cardiomyopathy, a Fibrotic and Inflammatory Form of Cardiomyopathy Distinct From Typical Dilated or Arrhythmogenic Right Ventricular Cardiomyopathy. Circulation 2020;141:1872–1884.

12. Carvajal-Huerta L. Epidermolytic palmoplantar keratoderma with woolly hair and dilated cardiomyopathy. J Am Acad Dermatol 1998;39:418–21.

13. Green KJ, Stappenbeck TS, Parry DA, Virata ML. Structure of desmoplakin and its association with intermediate filaments. J Dermatol 1992;19:765–9.

14. Sen-Chowdhry S, Syrris P, Ward D, Asimaki A, Sevdalis E, McKenna WJ. Clinical and genetic characterization of families with arrhythmogenic right ventricular dysplasia/cardiomyopathy provides novel insights into patterns of disease expression. Circulation 2007;115:1710–20.

15. Uzumcu A, Norgett EE, Dindar A et al. Loss of desmoplakin isoform I causes early onset cardiomyopathy and heart failure in a Naxos-like syndrome. J Med Genet 2006;43:e5.

16. O’Keefe EJ, Erickson HP, Bennett V. Desmoplakin I and desmoplakin II. Purification and characterization. J Biol Chem 1989;264:8310–8.

17. Landrum MJ, Lee JM, Benson M et al. ClinVar: public archive of interpretations of clinically relevant variants. Nucleic Acids Res 2016;44:D862–8.

18. Karczewski KJ, Francioli LC, Tiao G et al. The mutational constraint spectrum quantified from variation in 141,456 humans. Nature 2020;581:434–443.

19. Richards S, Aziz N, Bale S et al. Standards and guidelines for the interpretation of sequence variants: a joint consensus recommendation of the American College of Medical Genetics and Genomics and the Association for Molecular Pathology. Genet Med 2015;17:405–424.

20. Karczewski KJ, Francioli LC, Tiao G et al. Variation across 141,456 human exomes and genomes reveals the spectrum of loss-of-function intolerance across human protein-coding genes. bioRxiv 2019.

21. Bhonsale A, James CA, Tichnell C et al. Risk stratification in arrhythmogenic right ventricular dysplasia/cardiomyopathy-associated desmosomal mutation carriers. Circ Arrhythm Electrophysiol 2013;6:569–78.

22. Gomes J, Finlay M, Ahmed AK et al. Electrophysiological abnormalities precede overt structural changes in arrhythmogenic right ventricular cardiomyopathy due to mutations in desmoplakin-A combined murine and human study. Eur Heart J 2012;33:1942–53.

23. Ingles J, Bagnall RD, Yeates L et al. Concealed Arrhythmogenic Right Ventricular Cardiomyopathy in Sudden Unexplained Cardiac Death Events. Circ Genom Prec Med 2018;11:e002355.

24. Cheung CC, Davies B, Krahn AD. Letter by Cheung et al Regarding Article, “Concealed Arrhythmogenic Right Ventricular Cardiomyopathy in Sudden Unexplained Cardiac Death Events”. Circ Genom Precis Med 2019;12:e002447.

25. Cadrin-Tourigny J, Bosman LP, Nozza A et al. A new prediction model for ventricular arrhythmias in arrhythmogenic right ventricular cardiomyopathy. Eur Heart J 2019;40:1850–1858.

26. Rampazzo A, Nava A, Malacrida S et al. Mutation in human desmoplakin domain binding to plakoglobin causes a dominant form of arrhythmogenic right ventricular cardiomyopathy. Am J Hum Genet 2002;71:1200–1206.

27. Bauce B, Frigo G, Marcus FI et al. Comparison of clinical features of arrhythmogenic right ventricular cardiomyopathy in men versus women. Am J Cardiol 2008;102:1252–7.

28. Fairweather D, Cooper LT, Jr., Blauwet LA. Sex and gender differences in myocarditis and dilated cardiomyopathy. Curr Probl Cardiol 2013;38:7–46.

29. Ho CY, Day SM, Ashley EA et al. Genotype and Lifetime Burden of Disease in Hypertrophic Cardiomyopathy: Insights from the Sarcomeric Human Cardiomyopathy Registry (SHaRe). Circulation 2018;138:1387–1398.

30. Augusto JB, Eiros R, Nakou E et al. Dilated cardiomyopathy and arrhythmogenic left ventricular cardiomyopathy: a comprehensive genotype-imaging phenotype study. Eur Heart J Cardiovasc Imaging 2020;21:326–336.

31. Ingles J, Burns C, Bagnall RD et al. Non-familial Hypertrophic Cardiomyopathy: Prevalence, Natural History, and Clinical Implications. Circ Cardiovasc Genet 2017;10.

32. Ingles J, Goldstein J, Thaxton C et al. Evaluating the Clinical Validity of Hypertrophic Cardiomyopathy Genes. Circ Genom Precis Med 2019;12:e002460.

33. Maruthappu T, Posafalvi A, Castelletti S et al. Loss-of-function desmoplakin I and II mutations underlie dominant arrhythmogenic cardiomyopathy with a hair and skin phenotype. Br J Dermatol 2019;180:1114–1122.

34. Kapplinger JD, Landstrom AP, Salisbury BA et al. Distinguishing arrhythmogenic right ventricular cardiomyopathy/dysplasia-associated mutations from background genetic noise. J Am Coll Cardiol 2011;57:2317–27.

35. Rasmussen TB, Hansen J, Nissen PH et al. Protein expression studies of desmoplakin mutations in cardiomyopathy patients reveal different molecular disease mechanisms. Clin Genet 2013;84:20–30.

36. Bolling MC, Veenstra MJ, Jonkman MF et al. Lethal acantholytic epidermolysis bullosa due to a novel homozygous deletion in DSP: expanding the phenotype and implications for desmoplakin function in skin and heart. Br J Dermatol 2010;162:1388–94.

