## Supplementary Material for "Variant location is a novel risk factor for individuals with arrhythmogenic cardiomyopathy due to a desmoplakin (*DSP*) truncating variant"

Hoorntje ET*, Burns C* *et al.*

**SUPPLEMENTAL DATA**

**METHODS**

**Patient Cohort**

An international cohort of unrelated patients shown to have a *DSP*tv was assembled. The proband was defined as the first affected in the family who underwent genetic testing with a *DSP*tv identified. Cases seen in the specialised multidisciplinary Genetic Heart Disease clinic at Royal Prince Alfred Hospital between 2002-2018 were included. International centers were included via existing collaborative networks. These included the Department of Medical Genetics, University Medical Center Groningen, Groningen, The Netherlands; Department of Medical Genetics, University Medical Center Utrecht, Utrecht, The Netherlands; Department of Medical Genetics, Maastricht University Medical Center, Maastricht, The Netherlands; Department of Medical Genetics, Amsterdam University Medical Center, Amsterdam, The Netherlands; Department of Medical Genetics, Leiden University Medical Center, Leiden, The Netherlands; Department of Medical Genetics, Erasmus Medical Center, Rotterdam, The Netherlands; Department of Medical Genetics, Radboud University Medical Center, Nijmegen, The Netherlands; Stanford Inherited Cardiovascular Diseases Group, Stanford University, California USA; and Cardiovascular Research Centre, Royal Brompton & Harefield Hospitals London UK. Members of the Australian Cardiac Genetic Testing network (ACGT) were invited to contribute probands*.* The ACGT network includes cardiac genetic professionals throughout Australia, including >90 clinicians, scientists and genetic counsellors working towards a standardised cardiac genetic testing pathway. Members were contacted from July 2016 and invited to contribute cases until August 2018. All aspects of the study were performed according to institutional human research ethics committee approval.

**Eligibility Criteria**

The definition of *DSP*tv included any variant that was likely to reduce or abolish the *DSP* protein compared with the wildtype allele. Variant types included in this definition were frameshift, splice-site, nonsense and insertion or deletion variants. All variant nomenclature adhered to the Human Genome Variation Society sequence variant nomenclature recommendations. A frameshift variant was defined as a deletion, duplication or insertion involving a number of base pairs not divisible by three and therefore disrupting the triplet reading frame of the normal protein product. Splice-site variants were defined as variants affecting the donor and acceptor splice regions (±1, 2, 3, 4, 5) between the intron and exon boundary with the potential to disrupt messenger ribonucleic acid and alter the protein product. Nonsense variants were defined as a premature stop codon being introduced. Deletions and insertions were defined as a sequence change whereby one or more nucleotide was not present when compared with a reference standard and resulting in disruption to the reading frame leading to a premature truncation. Probands were included regardless of cardiac phenotype if they harboured one of the variants described above. Both deceased and living patients were included. Genetic testing was performed by the referring institution. This included both clinical grade sequencing by commercial and University accredited testing laboratories, and research-based genetic testing. Genetic testing was performed for clinical purposes and the approach evolved over time, including exome sequencing, or targeted and comprehensive gene panels performed by Sanger or next-generation sequencing.

**Genetic variant classification**

Disease variants were classified using the American College of Medical Genetics and Genomics and Association for Molecular Pathology (ACMG/AMP) standards for variant classification (1). In brief, the criterion pathogenic moderate (PM2) were allocated to variants with a frequency in the Genome Aggregation Database (gnomAD) of <0.04% (2), and pathogenic very strong (PVS1) was used for variants in the NMD competent regions. PVS1_strong was allocated to *DSP*tv in the constitutive NMD incompetent region. Use of PVS1 or PVS1_strong was used based on:

1. *DSP* is intolerant to LOF variation, with only 18% of expected LOF variants observed (LOF observed/expected score; oe 0.18), and LOF observed/expected upper bound fraction (LOEUF) of 0.26 based on gnomAD v2.1 (3).
2. *DSP*tv variants are enriched in DCM cases compared to controls, with significant enrichment in two DCM cohorts (4).
3. *DSP*tv in the constitutive NMD incompetent region are predicted to escape NMD, however functional evidence supports truncation or alteration of this domain as having a critical impact on the protein function, hence PVS1_strong was applied (5). Further, re-analysis of variants reported by Mazzarotto et al. (4) was performed based on gene region, with significant excess seen for all three regions: constitutive NMD competent (OR 37, 95%CI 16-90, p<0.0001), non-constitutive NMD competent (OR 25, 95%CI 9-73, p<0.0001) and constitutive NMD incompetent (OR 11, 95% CI 4-31, p=0.0006) compared to gnomAD v2.1 controls. There was a high etiologic fraction for all regions: 0.97, 0.96 and 0.91 respectively.

**Clinical assessment**

Clinical data from all patients (probands and family members) with a pathogenic or likely pathogenic *DSP*tv were collected retrospectively. Clinical information was obtained by review of the medical record and cardiac investigations from the referring institution. Review of the genetic result, ECG, transthoracic echocardiogram (TTE), 24-hour ambulatory ECG (Holter) monitoring, cardiac magnetic resonance (CMR) imaging, three-generation pedigree, postmortem report and correspondence from the treating geneticist and/or cardiologist were reviewed by the study team where possible.

**Diagnosis and clinical definitions**

The LV was considered to be involved when one or more of the following was present: LV ejection fraction <55%, presence of LV late gadolinium enhancement (LGE) or intramyocardial fat (including septum) on CMR imaging or pathologic abnormalities found on autopsy after sudden cardiac death (SCD). Cutaneous abnormalities were defined as palmoplantar keratoderma and/or woolly hair. Cutaneous abnormalities were not systematically investigated, but included if noted in the medical record. Regional wall motion abnormalities included those reported in either the right or left ventricle. Premature ventricular contractions were defined as >500/24 hours. Those with a primary ventricular arrhythmia phenotype had a high burden of premature ventricular complexes on Holter monitoring (>10% over 24 hours) or resuscitated cardiac arrest in the absence of structural and functional abnormalities of the myocardium assessed at cardiac evaluation. A composite outcome of ventricular arrhythmia included SCD, resuscitated cardiac arrest, appropriate implantable cardioverter-defibrillator (ICD) therapy, or sustained ventricular tachycardia. Appropriate ICD therapy was defined as anti-tachycardia pacing, or an ICD discharge for termination of ventricular tachycardia or fibrillation. SCD in probands was defined as sudden death in an otherwise healthy individual of any age within 1 hour after the onset of symptoms, or when unwitnessed, within 24 hours after the individual was last seen in good health. Family history of SCD included those with a suspicious death of a first-degree relative aged less than 40 years.

**Literature review**

We conducted a literature review of all *DSP*tv reported in PubMed (accessed on June 3, 2019). We selected *DSP*tv reported in publications through a search using the terms “*DSP*” or “desmoplakin” in combination with “mutation” or “variant” in title and/or abstract. No limitations were placed on language, type, or date of publications. This selection was matched with variants reported in HGMD Pro and the ARVC database (6). We selected all variants in patients with available clinical information. Particular care was taken in reviewing the cardiologic and ectodermal manifestations, age of patients at evaluation and diagnosis, additional performed genetic tests, family history and cascade screening, immunohistochemical, ultrastructural or functional analysis.

**RESULTS**

**Literature review of reported *DSP*tv**

Isolated cardiomyopathy, cardiocutaneous disease, and isolated cutaneous disorders were associated with both mono- and bi-allelic *DSP*tv. Cardiac manifestations reported included DCM, ACM (either right, left or biventricular involvement), SCD, early onset cardiac failure, and peripartum cardiomyopathy. Cutaneous abnormalities were congenital or appeared later, most commonly in the first year of life, and included palmoplantar keratoderma, woolly hair, hypotrichosis, alopecia, abnormally shaped teeth, enamel defects in both the deciduous and permanent dentition, tong erosion, nail dystrophy, and lethal acantholytic epidermolysis bullosa, with more severe manifestations seen in bi-allelic disease. Affected patients harboring homozygous or compound heterozygous *DSP*tv always presented with skin abnormalities, more often in association with an early onset cardiomyopathy (15 [75%] families) than as an isolated trait (5 [25%] families). As many patients without reported cardiomyopathy were evaluated in their first decade, a later-onset cardiomyopathy could not be ruled out in these families. When families were ascertained via a proband <10 years of age, early onset cardiomyopathy associated with cutaneous disease was the most prevalent phenotype, and disease was caused by biallelic *DSP*tv.

**Supplementary Figure 1: Overview of the systematic review search criteria**

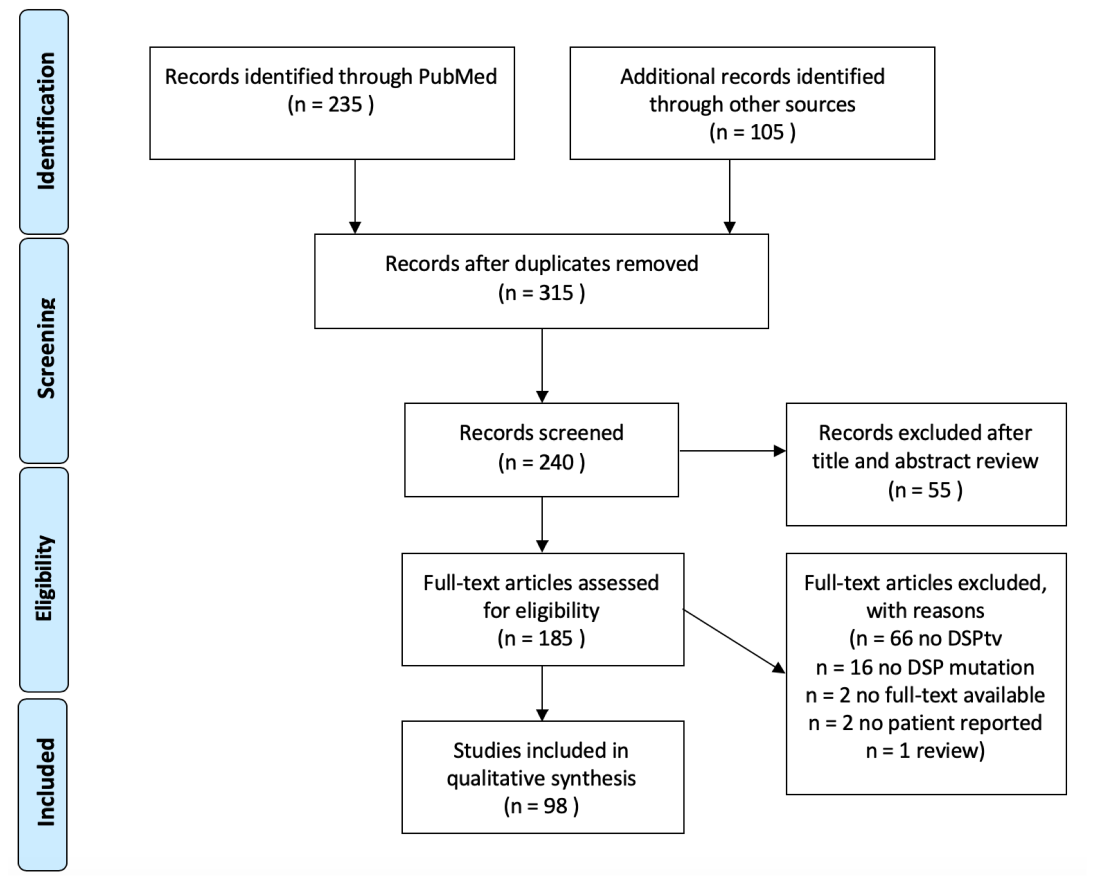

**References**

1.	Richards S, Aziz N, Bale S et al. Standards and guidelines for the interpretation of sequence variants: a joint consensus recommendation of the American College of Medical Genetics and Genomics and the Association for Molecular Pathology. Genetics in Medicine 2015;17:405-424.

2.	Karczewski KJ, Francioli LC, Tiao G et al. Variation across 141,456 human exomes and genomes reveals the spectrum of loss-of-function intolerance across human protein-coding genes. bioRxiv 2019.

3.	Karczewski KJ, Francioli LC, Tiao G et al. The mutational constraint spectrum quantified from variation in 141,456 humans. Nature 2020;581:434-443.

4.	Mazzarotto F, Tayal U, Buchan RJ et al. Reevaluating the Genetic Contribution of Monogenic Dilated Cardiomyopathy. Circulation 2020;141:387-398.

5.	Abou Tayoun AN, Pesaran T, DiStefano MT et al. Recommendations for interpreting the loss of function PVS1 ACMG/AMP variant criterion. Hum Mutat 2018;39:1517-1524.

6.	Lazzarini E, Jongbloed JD, Pilichou K et al. The ARVD/C genetic variants database: 2014 update. Hum Mutat 2015;36:403-10.

7.	Maruthappu T, Posafalvi A, Castelletti S et al. Loss-of-function desmoplakin I and II mutations underlie dominant arrhythmogenic cardiomyopathy with a hair and skin phenotype. Br J Dermatol 2019;180:1114-1122.

8.	Lopez-Ayala JM, Gomez-Milanes I, Sanchez Munoz JJ et al. Desmoplakin truncations and arrhythmogenic left ventricular cardiomyopathy: characterizing a phenotype. Europace : European pacing, arrhythmias, and cardiac electrophysiology : journal of the working groups on cardiac pacing, arrhythmias, and cardiac cellular electrophysiology of the European Society of Cardiology 2014;16:1838-46.
